# Genomic epidemiology and transmission dynamics of plasmids carrying New Delhi metallo-β-lactamase (*bla*_NDM_) at a single hospital system over five years

**DOI:** 10.64898/2026.05.14.26353212

**Authors:** Nathan J. Raabe, Emma G. Mills, Sarika Bapat, Marissa P. Griffith, Kathleen Shutt, Kady D. Waggle, Alexander J. Sundermann, Ryan K. Shields, Lora L. Pless, Graham M. Snyder, Lee H. Harrison, Daria Van Tyne

## Abstract

**Background:** Conjugative plasmids encoding New Delhi metallo-β-lactamase (*bla*_NDM_) pose a threat for the spread of carbapenem resistance among healthcare-acquired pathogens. Plasmid-associated outbreaks of *bla*_NDM_-producing bacteria can involve multiple bacterial species and persist over long time-periods, making their detection and control difficult. We systematically studied the genomic epidemiology of *bla*_NDM_-encoding plasmids detected within a single hospital system over a five-year period.

**Methods:** *bla*_NDM_-producing isolates were collected from clinical cultures as part of the Enhanced Detection System for Healthcare-Associated Transmission (EDS-HAT) genomic sequencing active surveillance program, or during infection prevention and control (IP&C) investigations. Isolates were identified as *bla*_NDM_ producers by polymerase chain reaction (PCR); the presence of plasmid-encoded *bla*_NDM_ genes was confirmed by sequencing on both Illumina and Oxford Nanopore platforms. Plasmids were clustered using Pling and bacterial relatedness of host isolates was evaluated with split kmer analysis. Electronic health record data were used to identify shared unit-level spatiotemporal exposures and epidemiologic links within both plasmid and host clusters.

**Results:** We identified 61 *bla*_NDM_-producing isolates collected from 54 patients sampled between November 2020 and July 2025. Isolates belonged to 15 Enterobacterales species; *Enterobacter hormaechei* was the most frequently sampled species (n=23, 37%), and *bla*_NDM-5_ was the most frequently observed *bla*_NDM_ allele (n=36, 59%). We observed six clusters of genetically similar *bla*_NDM_-encoding plasmids each containing 2–28 isolates, and eight singleton plasmids. The two largest plasmid clusters consisted of a highly conserved 46 kb IncX3 family *bla*_NDM-5_-encoding plasmid (n=28 plasmids, 9 species) and a more variable 98–201 kb IncC family *bla*_NDM-1_-encoding plasmid (n=12 plasmids, 6 species). Epidemiologic investigation paired with whole genome sequencing identified spatiotemporal associations between shared patient exposures and putative plasmid and bacterial transmission clusters, suggesting that unit-level exposures contribute to plasmid dissemination. Finally, analysis of publicly available sequences showed that the most prevalent plasmids detected, IncX3(*bla*_NDM-5_) and IncC(*bla*_NDM-1_), also demonstrated high global prevalence.

**Conclusions:** This study demonstrates the diversity of *bla*_NDM_ carrying plasmids within a single hospital system and their capacity to cause prolonged, multispecies outbreaks. Integrating whole genome sequencing with epidemiologic data identified unit-level spatiotemporal overlap as a likely contributor to plasmid dissemination in the hospital.

## BACKGROUND

The persistent burden of multidrug-resistant bacteria within hospital systems is, in part, driven by the continual propagation of antibiotic resistance-encoding plasmids (1). Plasmids are small, circular pieces of extrachromosomal DNA which can encode genes conferring antibiotic resistance to common antimicrobial treatments, enabling the bacteria carrying them to adapt to environments with increased antibiotic pressures, such as healthcare settings. Plasmids encoding antimicrobial resistance genes are primarily spread through either *bacterial transmission*, referring to transmission of the same plasmid-harboring bacterial species between patients, or through *plasmid transmission*, which refers to the spread of plasmids between the same or different bacterial species or lineages.

Due to the potential for inter-species or inter-lineage plasmid transmission, it can be difficult to detect, track, and intervene on plasmid transmission events. Currently, the spread of plasmid-associated resistance is not captured by most whole genome sequencing (WGS) surveillance systems designed to detect single-strain outbreaks driven by bacterial transmission (2–5). Thus, an approach using WGS to characterize both bacterial and plasmid transmission, alongside the incorporation of systematic epidemiologic investigation, is needed to detect and control plasmid-associated outbreaks (6).

The New Delhi metallo-β-lactamase (*bla*_NDM_, NDM) enzyme confers resistance to nearly all β-lactams, including the carbapenems, and is increasingly prevalent in the United States (7). Previously, we detected and investigated a multispecies IncX3 family plasmid-associated outbreak of *bla*_NDM-5_ infections at our hospital using enhanced genomic surveillance (8). Due to its clinical significance, continued *bla*_NDM_ surveillance beyond the initial outbreak investigation led to further detection of both *bla*_NDM-1_ and *bla*_NDM-5_ allelic variants, which were found to be associated with a variety of plasmid backbones. In this study, we aimed to characterize the dissemination and diversity of *bla*_NDM_-encoding plasmids identified within our healthcare center during a five-year period from 2020-2025.

## METHODS

### Study setting and isolate collection

Study isolates were recovered from patient cultures identified as NDM-producers obtained between November 2020 and July 2025 across five UPMC hospitals in the greater Pittsburgh region. Cultures were collected for either clinical care purposes as part of the Enhanced Detection System for Healthcare-Associated Transmission (EDS-HAT), by the UPMC XDR Pathogens Lab, or through infection prevention and control (IP&C) active surveillance of carbapenem-resistant Enterobacterales (CRE), as previously described (8). Briefly, active surveillance for detection of colonization was conducted for potentially exposed patients, typically defined by concomitant stay (for ≥1 day) on the same inpatient unit as a patient testing positive for a *bla*_NDM_-producing isolate, or within 3 days of the positive patient’s transfer. Active surveillance was performed using peri-anal swab screening, with at least one round performed during or after exposure and additional rounds performed at IP&C discretion based on the index patient’s length of stay, clinical condition, and prior surveillance results. Additional *bla*_NDM_-positive cultures from the same patient were excluded if the isolate was the same species and sequence type as a prior culture from that patient.

### Microbiologic methods

Isolates were sub-cultured onto blood agar plates for species identification using MALDI-TOF. Antimicrobial susceptibility testing was performed using the MicroScan WalkAway system (Beckman Coulter, Brea, CA, USA), and results were interpreted according to Clinical and Laboratory Standards Institute (CLSI) guidelines (9). Carbapenemase production was confirmed using the modified carbapenem inactivation test, performed as described in the CLSI Performance Standards for Antimicrobial Susceptibility Testing (M100) document (9). The presence of NDM was initially confirmed by either PCR for *bla*_NDM_ performed in a research laboratory or, beginning in May 2022, using the NG-Test CARBA 5 lateral flow assay performed in the clinical microbiology laboratory (Hardy Diagnostics, Santa Maria, CA).

### Whole genome sequencing

Isolates were streaked onto a blood agar plate (BD, Franklin Lakes, NJ) and grown overnight at 37°C; DNA was extracted from single bacterial colonies using the MagMax DNA Multi-Sample Ultra 2.0 extraction kit on a KingFisher Apex (Thermo Fisher Scientific) or using a DNeasy Blood and Tissue Kit (Qiagen, Hilden, Germany). Both short- and long-read whole genome sequencing were performed for each study isolate. Short-read sequencing was performed using the Illumina MiSeq, NextSeq 550 or 1000, or NovaSeq X Plus platform (Illumina, San Diego, CA). Libraries were prepared using an Illumina DNA Prep (M) Tagmentation Kit on an EpMotion, sequenced (v2.5 300-cycle kit) using paired-end reads (2 × 300 bp on MiSeq; 2 × 150 bp on NextSeq and NovaSeq platforms), and demultiplexed using bcl2fastq v2.20. Long-read sequencing was performed using an Oxford Nanopore Technologies MinION Mk1C device with R9.4.1 or R10.4.1 flow cells and SQK-RBK004 or SQK-RBK114.24 rapid gDNA barcoding kits. Basecalling and demultiplexing were performed using Albacore v2.3.3 (default parameters), Guppy v2.3.1/v6.3.9 (default parameters), or Dorado v0.9.6 (high accuracy model; dna_r10.4.1_e8.2_400bps_hac@v5.0.0).

### Genome sequencing quality control, assembly, and strain typing

Initial short-read genome assemblies were constructed for all isolates using SPAdes v3.15.5 (10) and genomes were annotated with Prokka v1.14.5 (11). Bacterial species were determined by comparisons with typed strains of suspected species using fastANI (12). Short-read assembly quality was assessed using QUAST v5.2.0 (13); isolates with estimated mean Illumina sequencing depth <35x were excluded from further analyses. Multilocus sequence types (STs) were determined using PubMLST (https://github.com/tseemann/mlst) and the PubMLST database (14). For optimal characterization of plasmids, hybrid assemblies were constructed from both Illumina and Nanopore sequencing data for all study isolates using Unicycler v0.5.0 (15). Long-read sequencing was performed with a target depth of ≥10x, and for some isolates was repeated to obtain sufficient depth to resolve complete circular plasmid sequences from hybrid assembled genomes.

### Plasmid identification and typing

Study plasmids were identified as circular contigs from hybrid assemblies encoding identifiable plasmid replicons and a *bla*_NDM_ allele (except for three contigs which could not be circularized). Incompatibility and replicon types were assigned using MOB-typer (v3.1.9) (16); replicon profiles were confirmed against replication machinery gene hits detected by BLASTn against the PlasmidFinder database (≥80% similarity; [% identity * % coverage] / 100) (17) (**Additional file 1: Table S1**). For each plasmid, *bla*_NDM_ allele type was determined via hits detected by BLASTn against the AMRFinderPlus database (≥100% similarity) (18).Throughout the text, study plasmids are described by their combination of replicon type profile and *bla*_NDM_ variant (e.g., IncC/IncFIB(*bla*_NDM-1_)).

### Plasmid similarity and clustering

Plasmids were clustered using Pling v2.0.0, applying stringent thresholds recommended for epidemiologic investigations (containment distance <0.3; DCJ-Indel distance ≤4) to define clusters of highly similar plasmids **(Additional file 2: Fig. S1)** (19). If the smaller plasmid in a pairwise comparison was ≥120 kb, we allowed +1 DCJ for each 20 kb range above this threshold. Each cluster containing >1 plasmid was assigned a numeric label in descending size order (*i.e*., C1–C6), with ties broken by earliest isolate culture date. Plasmids not meeting the criteria for inclusion into a cluster were labeled as singletons. Plasmid study identifiers (*i.e*., p1–p61) were assigned sequentially beginning with plasmids from the largest plasmid cluster. Within each cluster, plasmids were numbered in chronological order by host isolate culture date. For direct comparisons made between IncC and IncC/IncFIB plasmids from plasmid cluster C2, annotation was performed using Bakta v1.5.1 (20) and plasmids were aligned and visualized using EasyFig v2.2.2 (21).

### Bacterial host strain relatedness

To define bacterial host relatedness, pairwise single nucleotide polymorphism (SNP) distances were calculated between study isolates of the same species and MLST using split kmer analysis (SKA) v1.0 (22) performed on short-read sequencing data. Host bacterial transmission clusters were defined to discriminate plasmid sharing due to clonal expansion of host strains versus plasmid sharing due to plasmid transmission. The threshold for host strain relatedness (*i.e.*, same host bacterial cluster) was defined as ≤ 30 SNPs using single linkage clustering, and was chosen based on the empirical distribution of within-sequence type pairwise SNPs observed between study isolates (**Additional file 2: Fig. S1**). Bacterial host clusters were named within plasmid clusters by size, with ties broken chronologically by culture date (*e.g.*, the largest host bacterial cluster within plasmid cluster C1 was designated “C1b1”). For any two isolates within a bacterial host cluster, we attributed the presence of highly similar plasmids to bacterial transmission rather than plasmid transmission via horizontal gene transfer.

### Epidemiologic data collection and statistical analysis

Temporal trends in the frequency of *bla*_NDM_-encoding plasmids recovered during the study period were assessed using negative binomial regression and 3-month intervals as the unit of analysis, starting at the earliest culture date, and included an offset term to account for the truncated final interval. The difference in plasmid-encoded *bla*_NDM_ frequency during study year 1 versus years 2–5 was assessed using a separate negative binomial regression model with a binary predictor. All statistical analyses were performed in R (v4.3.1). UPMC hospital stay data for each patient were obtained for the full five-year study period, beginning two months prior to the first isolate culture date (study day 1) and extending two months after the final culture date. For patients with missing stay data, the hospital where the culture was initially obtained was recorded. For stays with a missing start or end date, the stay was truncated to a single day. Hospital stay records were converted to within-unit stay pairs by linking each patient to a single plasmid (patient-plasmid entity), and comparing all eligible unit stays across entities (as defined below). Within-unit stay pairs were excluded if they involved the same patient-plasmid entity or distinct plasmids from the same patient. Patients in each stay pair were ordered temporally using the first positive culture date recorded for each patient, such that Patient A had the earlier culture date and Patient B had the later culture date. A temporal overlap window (±365 days before and after stay) was applied around the stay of Patient A. If the stay of Patient B intersected with the temporal overlap window of the stay of Patient A, this qualified as a shared-exposure stay pair (**Additional file 2: Fig. S2**). This window was intended to capture the full range of plausible exposure periods and reflects the variable reported durations of carbapenemase-producing Enterobacterales carriage, which are typically months but have been shown to persist beyond one year in some patients (23–28). Exposure pairs where overlap only occurred after both patients had tested positive did not qualify and were dropped. Qualifying shared-exposure stay pairs were retained and collapsed to the unit level, and measures of overlap timing, duration, and concordance between plasmid or host bacterial cluster were calculated (**Additional file 3: Extended Data File S1**). For each plasmid cluster and host bacterial cluster, unit-specific 2×2 tables were constructed to estimate the odds that a qualifying shared-exposure stay pair on that unit was cluster-concordant versus non-concordant, compared with pairs on all other units combined. Unit-level odds ratios were estimated using Fisher’s exact test with Haldane–Anscombe correction for zero cells where applicable, and the false discovery rate across tests was controlled using the Benjamini–Hochberg procedure.

### Global contextualization of locally circulating *bla*_NDM_-encoding plasmids

For each of the six defined plasmid clusters, the earliest isolated plasmid was selected and compared to the National Center for Biotechnology Information (NCBI) nucleotide database on November 4^th^, 2025 using BLASTn and capturing 1,000 hits per plasmid (29). NCBI plasmids were included in the study if they met the following criteria: i) BLASTn similarity score was ≥90% (≥99.99% for IncX3(*bla*_NDM-5_) plasmids due to high sequence conservation), ii) the presence of the same *bla*_NDM_ allele as observed in the respective plasmid cluster, and iii) at least one of the same plasmid replicons identified in the respective plasmid cluster. Phylogenetic trees of each plasmid cluster including both UPMC plasmids and NCBI plasmids meeting the above criteria were constructed based on a core gene alignment produced by Roary v3.13.0 (30) with genes annotated using Prokka v1.14.5 (11). Gaps and areas of high recombination (≥ 10 SNPs in a 100 bp sliding window) were removed from core genome alignments using Geneious v2023.1.1. RAxML-HPC v8.2.12 using the GTRGAMMA algorithm with 100 bootstraps was used to build phylogenetic trees from filtered core genome alignments (31). Phylogenetic trees and metadata were visualized using iTOL v7 (32).

## RESULTS

### Frequency of NDM-encoding plasmids at a single health system increased over time

In April 2022, the isolation of four *bla*_NDM_-producing carbapenem-resistant Enterobacterales (CRE) from the clinical cultures of three patients revealed a multispecies plasmid-associated outbreak of Enterobacterales isolates harboring an IncX3 plasmid encoding *bla*_NDM-5_ (8). This finding prompted both prospective genomic surveillance of CRE and a retrospective analysis of CRE isolates collected prior to April 2022. To understand the diversity and transmission dynamics of plasmids encoding *bla*_NDM_ at our center, we sequenced the genomes of all *bla*_NDM_-positive patient isolates collected between November 2020 and July 2025 from five UPMC hospitals located in the Pittsburgh, Pennsylvania area. Over this time, we collected 61 isolates with plasmid-encoded *bla*_NDM_ from 54 patients; five patients contributed two or more *bla*_NDM_-positive isolates from different Enterobacterales species. Plasmids carried one of two *bla*_NDM_ gene variants, including *bla*_NDM-1_ (n=25, 41%) and *bla*_NDM-5_ (n=36, 59%) (**Fig. 1A, Additional file 1: Table S1)**. Samples were most commonly isolated from rectal surveillance swabs (n=22, 36%), blood (n=15, 25%), and urine (n=14, 23%). Of the 15 different species observed, *Enterobacter hormaechei* was the most common (n=23, 38%), followed by *Klebsiella pneumoniae* (n=8, 13%), and *Escherichia coli* (n=6, 10%).

**Figure 1:**
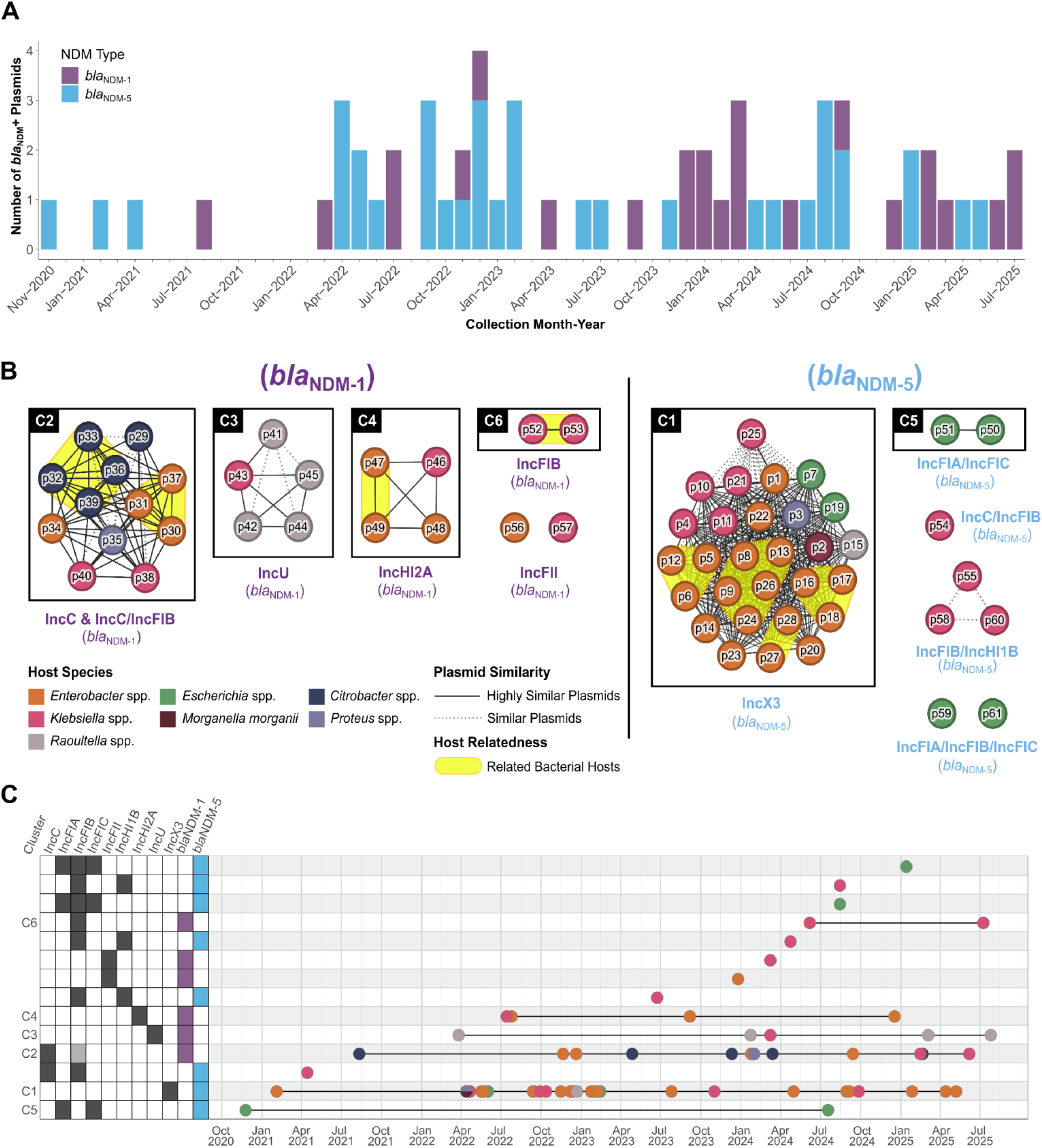
Incidence and genetic relatedness of *bla*_NDM_ plasmids at a single hospital over five years. **(A)** Temporal distribution of 61 *bla*_NDM_ plasmids, encoding either *bla*_NDM-1_ (purple) or *bla*_NDM-5_ (blue) observed in isolates from 54 patients collected between Nov 2020 and July 2025. **(B)** *bla*_NDM_-harboring plasmids clustered by genetic relatedness. Similar plasmids were defined as those with containment distance <0.3, and highly similar plasmids as those that also had a DCJ-Indel distance ≤4. Bacterial hosts separated by ≤30 SNPs were considered related. **(C)** Temporal dynamics of plasmid clusters. Plasmids were grouped by replicon profile and *bla*_NDM_ allele; cluster designations correspond to highly-similar plasmid clusters shown in (B). Dark grey: plasmid replicon gene present in all cluster plasmids; light grey: plasmid replicon gene present in some cluster plasmids; white: plasmid replicon gene absent. Plasmid nodes are colored by host species as in (B).

The frequency of isolates harboring plasmid-encoded *bla*_NDM-1_ or *bla*_NDM-5_ alleles observed at our healthcare facility throughout the study period were largely similar, ranging from 0–4 *bla*_NDM-1_–encoding plasmids and 0–5 *bla*_NDM-5_–encoding plasmids per 3-month interval, respectively. Early in the collection period (November 2020 to March 2022), the frequency of *bla*_NDM_-encoding plasmids was sporadic. However, between April 2022 and July 2025, we observed markedly higher frequencies of plasmid-encoded *bla*_NDM-1_ and *bla*_NDM-5_, consistent with the possibility of multiple plasmid-associated hospital outbreaks or increased importations. Overall, the frequency at which plasmid-encoded *bla*_NDM_ was observed per 3-month interval increased significantly over the study period (p = 0.041). Additionally, the frequency of plasmid-encoded bla_NDM_ per 3-month interval in years 2–5 was 3.9 times as high as that observed in year 1 (mean 3.8 vs. 1.0 isolates per interval; count ratio = 3.88, 95% CI 1.58–12.89, p = 0.009).

### Genetic relatedness of *bla*_NDM_-encoding plasmids identifies putative plasmid transmission clusters

We identified 11 distinct *bla*_NDM_-encoding plasmid types, as defined by replicon type profile and *bla*_NDM_ allele, ranging in length from 45 kb to 426 kb (**Additional file 1: Table S1**). Despite the diversity of *bla*_NDM_-carrying plasmid backbones observed, many were either IncX3(*bla*_NDM-5_) (n = 28, 46%) or IncC(*bla*_NDM-1_) (n = 10, 16%). Comparative genomic analysis identified six clusters of genetically related plasmids (clusters C1–C6; 2–28 plasmids per cluster, median=5 plasmids per cluster) and eight singleton plasmids, indicating the presence of both apparently isolated introductions and expansion of circulating plasmid types (**Fig. 1B**). Nested within the six plasmid clusters we identified nine bacterial host clusters. Bacterial host relatedness (≤ 30 SNPs) explained plasmid sharing in 13 of 28 (46.4%) isolates in plasmid cluster C1, 7 of 12 (58.3%) isolates in cluster C2, 2 of 4 (50.0%) isolates in cluster C4, and 2 of 2 (100%) isolates in cluster C6, but in none in clusters C3 (0 of 5, 0%) or C5 (0 of 2, 0%). Overall, bacterial transmission only accounted for 24/61 (39%) of clustered plasmids, suggesting that plasmid transmission contributed to plasmid spread independently from bacterial transmission.

Temporal analysis showed persistence of several plasmid clusters over multiple years **(Fig. 1C, Additional file: Table S1)**, consistent with ongoing circulation and/or repeated introductions of shared plasmid lineages. Persistence and frequency of detection varied by plasmid cluster. Plasmid cluster C1 (IncX3(*bla*_NDM-5_)) persisted for approximately 4.2 years and was frequently detected throughout the study period (median days between detection = 21). In contrast, cluster C2 (IncC and IncC/IncFIB(*bla*_NDM-1_) also persisted over a long interval (3.8 years) but was detected less frequently (median days between detection = 107). Other clusters persisted for 3–4 years, with less frequent detection, often separated by several month-long intervals. Taken together, these data suggest that bla*_NDM_*-encoding plasmid clusters may persist in healthcare settings for multiple years with varied frequencies of detection.

The largest plasmid cluster (cluster C1, plasmids p1–p28, February 2021 to January 2025) consisted of a highly conserved conjugative 46–73 kb IncX3(*bla*_NDM-5_) plasmid detected across nine species, consistent with extensive cross-species plasmid dissemination (8,33). Most plasmids in this cluster shared nearly all sequence content (mean containment distance between plasmids = 0.004) and had minimal structural differences (mean DCJ-Indel distance between plasmids = 0.5) **(Additional file 3: Extended Data File S1)**. Notably, the singular 73 kb plasmid in this cluster encoded additional resistance determinants, including the carbapenemase *bla*_OXA-181_ and the 16S rRNA methyltransferase *rmtB1*, an aminoglycoside resistance gene (**Fig. S3**) (34). Of the 28 plasmids in plasmid cluster C1, 13 (46.5%) were part of a host bacterial cluster (**Fig. 1B**). All four bacterial host clusters (C1b1–b4) consisted of *Enterobacter* spp. isolates (*E. hormaechei* ST45 and *E. roggenkampii* ST997). The remaining isolates in this plasmid cluster (n=15, 53.5%) showed only distant relatedness to other study isolates, indicating that independent introductions into the hospital, plasmid transmission, or a combination thereof likely played just a big a role as bacterial transmission in the dissemination of the IncX3(*bla*_NDM-5_) plasmid in our hospital system.

The second largest plasmid cluster (cluster C2, plasmids p29–p40, August 2021 to June 2025) was comprised of 12 structurally variable conjugative 98–201 kb IncC(*bla*_NDM-1_) and IncC/IncFIB(*bla*_NDM-1_) plasmids (avg. containment = 0.078; avg. DCJ = 3.2) encoded by isolates spanning six different species. In addition to *bla*_NDM-1_, most of these plasmids (n=8, 66%) carried the extended-spectrum beta-lactamase *bla*_CTX-M-14_, and three (25%) plasmids harbored the 16S rRNA methlytransferase *rmtC*. The majority of plasmids in this cluster encoded the IncC replicon alone (n=10, 83%), however, two plasmids (17%) encoded an additional IncFIB replicon (**Fig. 2A**). Plasmid cluster C2 contained two bacterial host clusters (C2b1 and C2b2), which consisted of four isolates belonging to *Citrobacter portucalensis* ST401 and three belonging to *E. hormaechei* ST78, respectively (**Fig. 1B**). Plasmid sequence comparisons within the four *C. portucalensis* ST401 isolates from bacterial host cluster C2b1, including two isolates harboring IncC(*bla*_NDM-1_) plasmids and two isolates harboring IncC/IncFIB(*bla*_NDM-1_) plasmids, revealed evidence of recombination resulting in plasmid fusion: the IncC(*bla*_NDM-1_) plasmid backbone common to cluster C2 was nearly fully contained within the 195–201 kb IncC/IncFIB(*bla*_NDM-1_) sequence observed in plasmids p33 and p36 (**Fig. 2A**, **Fig. 2B**). The two isolates in this host bacterial cluster with IncC(*bla*_NDM-1_) plasmids were each also found to contain a separate, circularized 78 kb IncFIB plasmid with >99.9% BLASTn similarity to plasmids p33 and p36 **(Additional file 3: Extended Data File S1).** Taken together, these results suggest recombination-driven plasmid evolution, where two plasmids appeared both fused and unfused in the assemblies of genetically related isolates over time.

**Figure 2:**
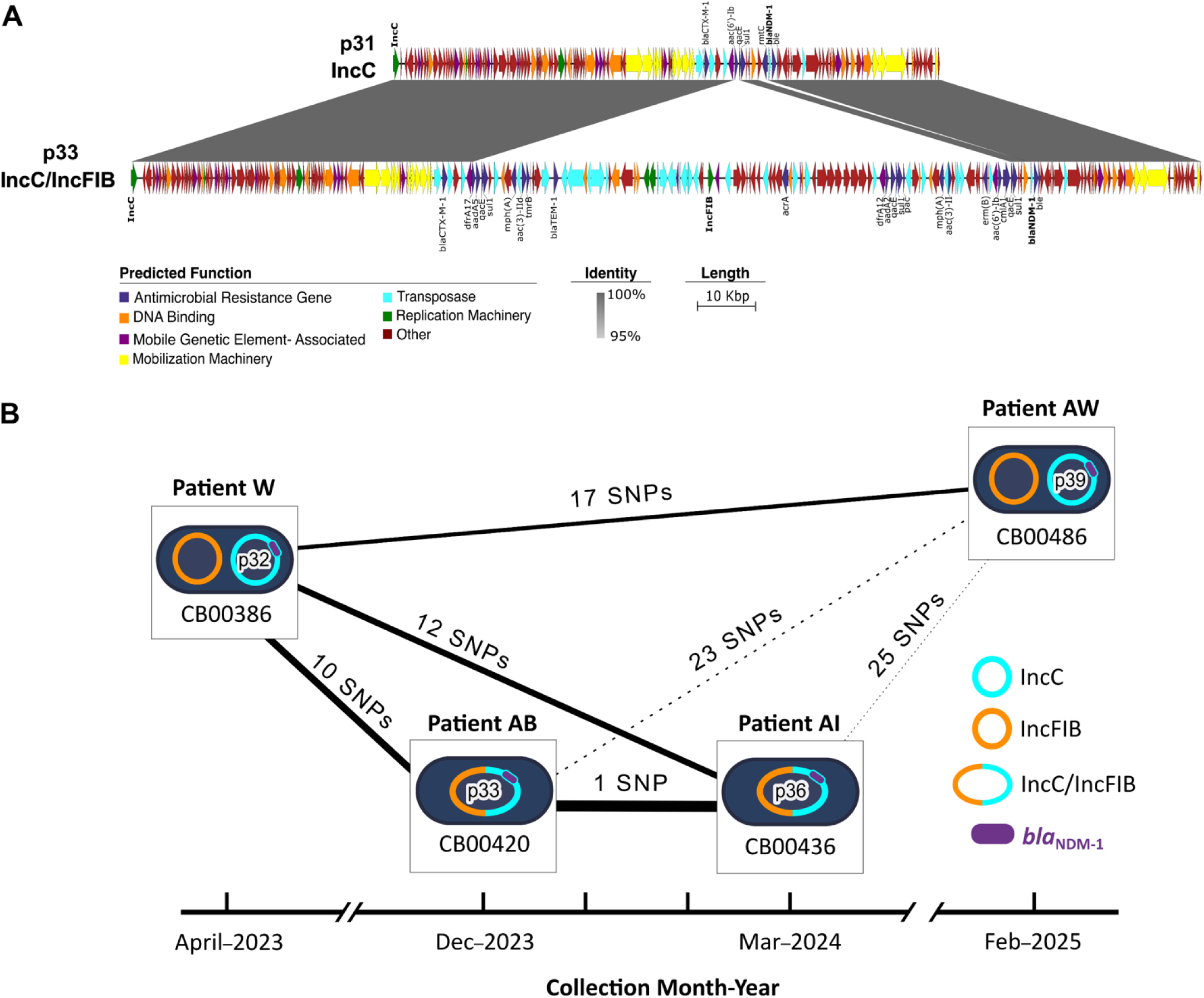
Sequence variability of representative IncC(*bla*_NDM-1_) and IncC/FIB(*bla*_NDM-1_) plasmids. **(A)** Nucleotide sequence alignment between cluster C2 plasmids p31 (IncC) and p33 (IncC/IncFIB) were visualized using EasyFig v2.2.2. Genes were annotated using Bakta v1.9.2 and were grouped into predicted functional categories as indicated. Nucleotide identity, calculated from BLASTn comparisons using a 1,000-bp window size, is shown from 95% (light grey) to 100% (dark grey). **(B)** IncC, IncFIB, and mosaic IncC/IncFIB plasmid replicon carriage is shown over time for the four related *Citrobacter portucalensis* isolates within bacterial cluster C2b1. Pairwise host isolate SNP differences are shown above each edge; thicker line weights indicate greater host relatedness (i.e., lower SNPs). Dashed lines are shown for connections with >20 SNPs. The presence of *bla*_NDM-1_ is shown as a purple segment on the corresponding plasmid replicon.

### Unit-level spatiotemporal associations are stronger for plasmid clusters than bacterial host clusters

A total of 3907 same-unit stay-pairs were observed across all patients, of which 1652 met criteria to be considered a qualifying shared-exposure stay-pair (i.e., overlapping unit stays ±365 days) **(Additional file 3: Extended Data File S1)**. These qualifying exposure stay-pairs were observed across 36 units from two of five hospitals, with virtually all pairs occurring at Hospital I (n = 1636, 99%) and a small fraction occurring at Hospital II (n = 16, 1%). Six patients (11%) were missing hospital stay data; however, the hospitals where cultures were obtained were known. Missingness was distributed without any discernable pattern across plasmid clusters, species, specimen sources, and study years **(Additional file 1: Table S1)**. We calculated unit-level odds ratios for both plasmid and host clusters based on qualifying shared-exposure stay pairs **(Fig. 3; Additional file 1: Table S2)**. There were 28 hospital units that, for at least one plasmid or bacterial cluster, had ≥1 qualifying within-cluster shared-exposure stay pair and an exposure odds ratio > 1. Of these, 14 were medical/surgical units (50.0%), nine were intensive care units (32.1%), two were post-operative care units (7.1%), two were emergency rooms (7.1%), and one was a rehabilitation unit (3.6%).

**Figure 3:**
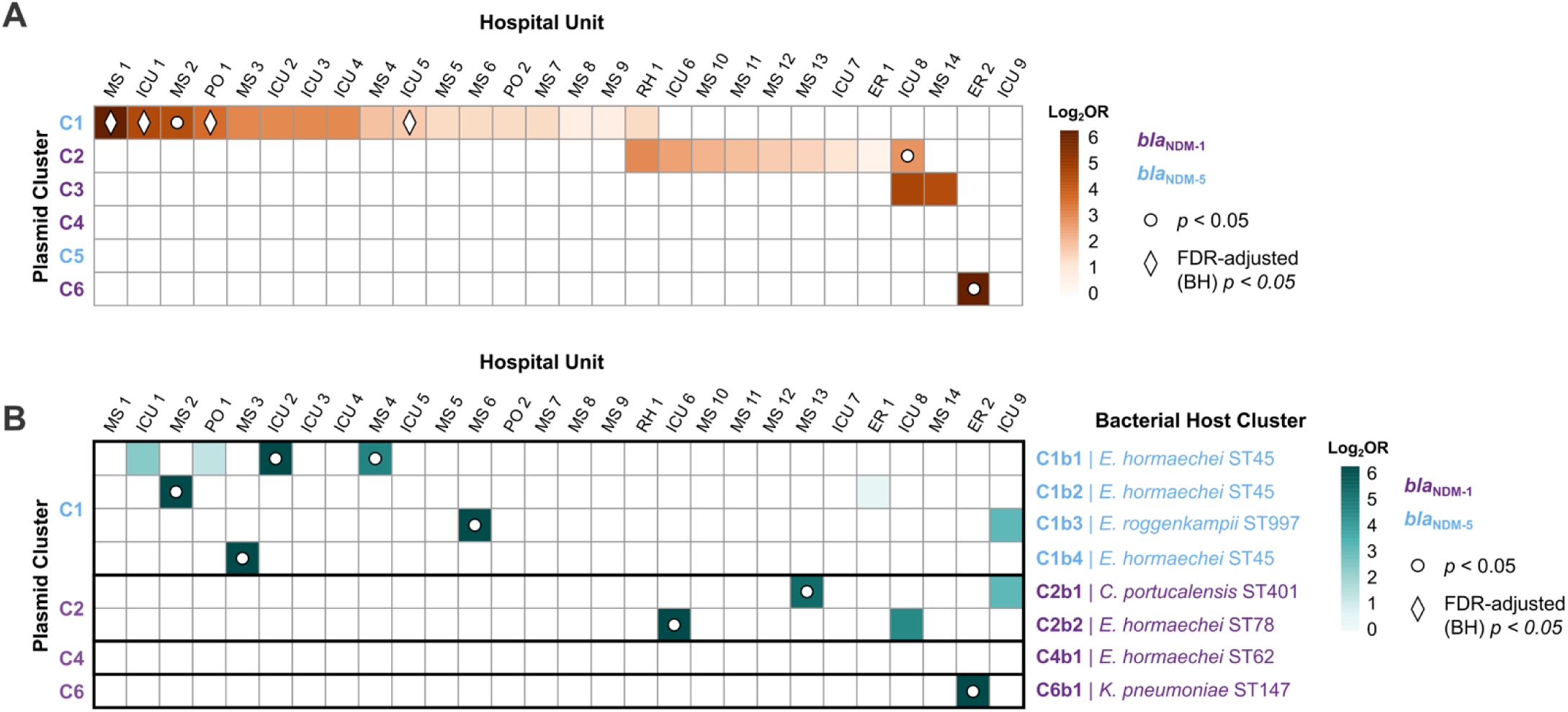
Unit-level spatiotemporal associations between shared exposures and plasmid or bacterial host clusters. Heatmaps show, for each hospital unit, the log₂ odds ratio (OR) that a qualifying shared-exposure stay pair involved two entities in the **(A)** same plasmid cluster, or **(B)** same host bacterial cluster, compared with all other units combined. Units with at least 1 qualifying stay pair for any plasmid or host cluster (colored by NDM-allele) are shown. Odds ratios were tested using Fisher’s exact test with Haldane–Anscombe correction for zero cells, and Benjamini–Hochberg (BH) adjustment was used to control for the false discovery rate (FDR) across tests. White dots denote unadjusted *p* < 0.05 and diamonds denote FDR-adjusted *p* < 0.05. Hospital units are annotated by type; MS, medical/surgical; ICU, intensive care unit; PO, post-operative; RH, rehabilitation; ER, emergency room.

Within-cluster overlap was concentrated on a subset of units for each plasmid cluster, as reflected by higher unit-specific odds that a qualifying shared-exposure stay pair on those units was cluster-concordant compared with pairs on all other units combined **(Fig. 3A; Additional file 1: Table S2)**. For example, in plasmid cluster C1, 17 units showed increased odds of within-cluster concordance, including several strong associations, such as those with an intensive care unit (ICU–1), medical/surgical unit (MS–1), and a post-operative care unit (PO–1) (OR range 11.6–65.1; BH-adjusted p < 0.001). Plasmid cluster C1 also showed associations with several other units, including a significant but weaker association with another intensive care unit (ICU-5, OR 3.3, BH-adjusted p < 0.001), and 14 other units with weaker odds ratios which did not reach statistical significance after correction for multiple hypothesis testing. Other plasmid clusters (C2, C3, C6) also showed associations with select ICU, rehabilitation, emergency, and medical/surgical units, with two associations reaching statistical significance before Benjamini–Hochberg correction (C2 with ICU-8, OR 6.8, p < 0.05; C6 with ER-2, OR 10.2, p < 0.01). In contrast, some units (*e.g.*, emergency room ER–1) contributed many qualifying shared-exposure stay pairs across multiple plasmid clusters, but as a result, clusters associated with this unit had odds ratios close to or less than one, indicating the within-cluster overlap experienced on these units was not elevated relative to overlap involving other clusters that occurred on the unit. While these types of units may initially appear to be important for plasmid transmission based on crude exposure counts within each cluster, their lack of specificity to any single cluster indicates that they instead represent a widespread exposure that is not specifically informative for understanding plasmid transmission.

Across bacterial host clusters, the units showing strong within-cluster associations were largely a subset of those with strong associations within their respective plasmid clusters. For example, all five associations across the four bacterial clusters (C1b1-C1b4) of plasmid cluster C1 also had within-plasmid cluster associations for the same units (**Fig. 3B; Additional file 1: Table S2**). This result was expected, as bacterial host clusters in this study were completely nested within plasmid clusters, and without overlap across clusters (**Fig. 1B**). Additionally, Cluster C6, where the presence of shared plasmids was driven entirely by bacterial transmission, had a single association to emergency room ER-2 for both plasmid and bacterial host clusters, indicating that this unit may have been an important route for bacterial transmission between patients. Notably, several units with strong within-plasmid cluster associations showed little to no host-cluster enrichment, suggesting that unit-level spatiotemporal associations for plasmid clusters are only partially explained by bacterial spread, and that spatiotemporal associations could be informative for understanding plasmid transmission across multiple bacterial hosts.

### Global circulation and conservation of locally disseminated plasmid backbones

To determine whether plasmids involved in putative transmission at our center represented globally circulating plasmids; we compared our local plasmids to a curated global collection. Briefly, for each local plasmid cluster, we used the first isolated plasmid as a reference and used it to query the NCBI database using BLASTn. From the BLASTn results, we selected plasmids with a similarity score ≥90% compared to the reference plasmid, presence of the same *bla*_NDM_ allele, and a similar plasmid replicon profile to the reference plasmid. Four of six plasmid clusters yielded hits meeting these inclusion criteria, and these were retained for downstream analysis. To investigate global diversity within these plasmid lineages, we constructed a phylogenetic tree from a core genome alignment of each plasmid. Consistent with their high local prevalence at UPMC, global IncX3(*bla*_NDM-5_) plasmids (n = 234) exhibited low genetic diversity and showed strong geographic enrichment, with the majority of plasmids isolated from Asia (77%) (**Fig. 4A; Additional file 1: Table S3**). In contrast, global IncC/IncFIB(*bla*_NDM-1_) plasmids (n = 63) displayed a broader geographic distribution across Asia (37%), Europe (19%), and North America (19%), yet remained highly conserved (**Fig. 4B**). Of concern, nearly all global IncC(*bla*_NDM-1_) plasmids (98%) additionally carried the 16S rRNA methyltransferase *rmtC*. Similar genetic conservation was observed among global IncFIB(*bla*_NDM-1_) plasmids (n = 25), which were isolated from three continents (**Fig. 4C**). In contrast, global IncFIA/IncFIC(*bla*_NDM-5_) plasmids (n = 20) showed greater sequence diversity, suggesting more frequent mutations and/or backbone recombination in this lineage (**Fig. 4D**).

**Figure 4:**
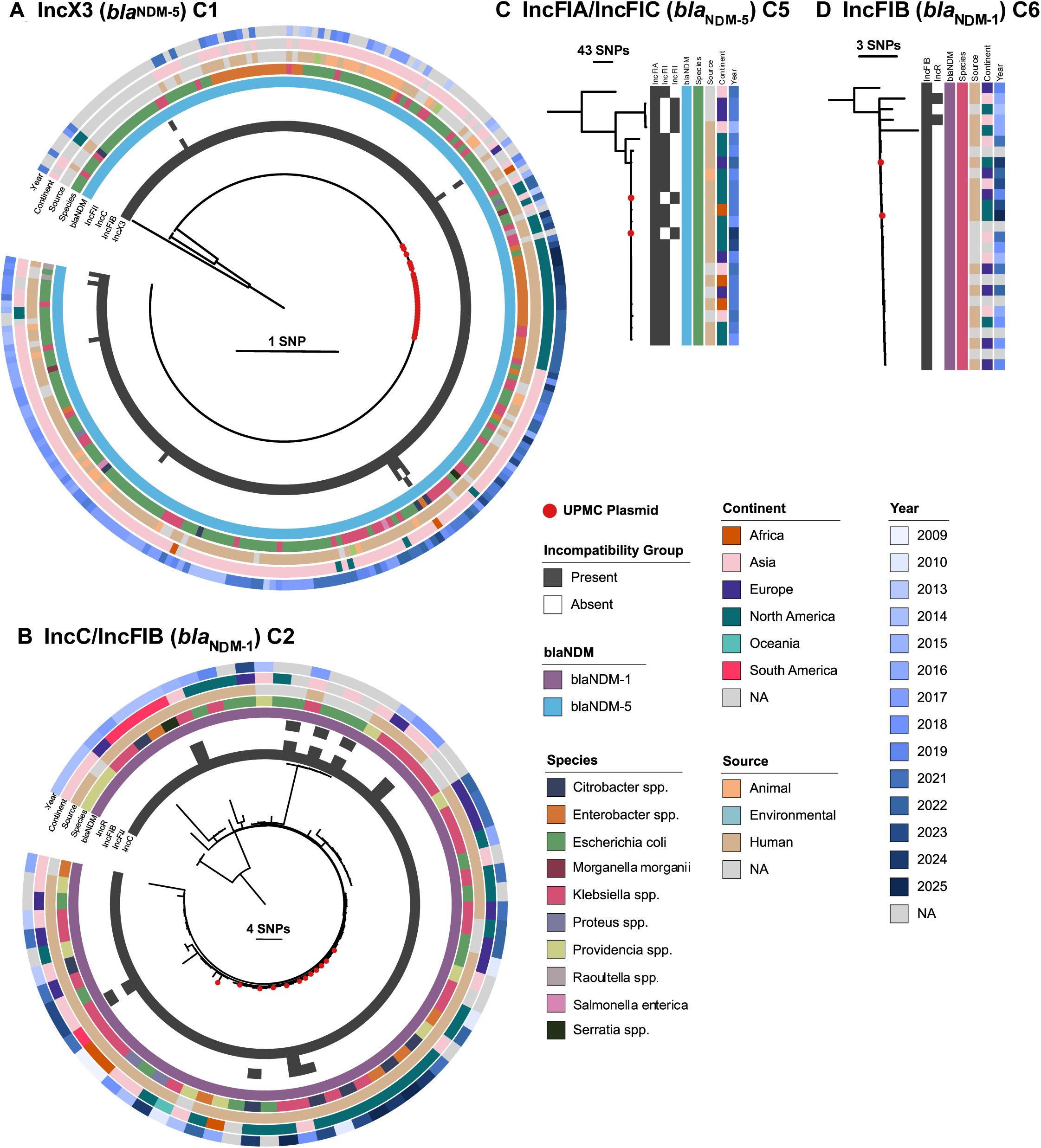
Global context of local *bla*_NDM_ plasmid clusters. Phylogenetic trees were constructed with RAxML based on plasmid core gene alignments produced by Roary. All trees are midpoint rooted. **(A)** IncX3(*bla*_NDM-5_) phylogeny based on 24 core genes shared between 262 plasmids. **(B)** IncC/IncFIB(*bla*_NDM-1_) phylogeny based on 99 core genes shared between 75 plasmids. **(C)** IncFIA/IncFII(*bla*_NDM-5_) phylogeny based on 79 core genes shared between 22 plasmids. **(D)** IncFIB(*bla*_NDM-1_) phylogeny based on 42 core genes shared between 27 plasmids. Local UPMC plasmids are indicated with red circles. Bacterial host species are colored as indicated. Available metadata (continent, isolation source, culture year) are shown; NA = not available.

Distinct associations between *bla*_NDM_-encoding plasmids and bacterial hosts were also observed. Of the 342 plasmids identified in our global analysis, the most frequent bacterial host was *E. coli* (n = 188, 55%) followed by *K. pneumoniae* (n = 91, 27%) and *E. hormaechei* (n = 16, 5%). While the IncX3(*bla*_NDM-5_) and IncC(*bla*_NDM-1_) plasmids were detected in several different species, the IncFIB(*bla*_NDM-1_) plasmid was only found in *K. pneumoniae* and the IncFIA/IncFIC(*bla*_NDM-5_) plasmid was only found in *E. coli* (**Fig. 4C**, **Fig. 4D**). Among the 243 global plasmids with available isolation source metadata, most were isolated from humans (n = 221, 87%), while animal-associated sources accounted for 12% (n = 29), predominantly among IncX3(*bla*_NDM-5_) plasmids. Temporal metadata of local and global combined plasmids suggested lineage-specific circulation dynamics. IncX3(*bla*_NDM-5_) plasmids, which were first identified in 2013, showed a marked increase in frequency starting in 2016 (**Additional file 2: Fig. S4A**). Similarly, the IncC/IncFIB(*bla*_NDM-1_) plasmid was first observed in 2009 and seems to have increased in frequency over the past decade (**Additional file 2: Fig. S4B**). IncFIA/IncFIC(*bla*_NDM-5_) and IncFIB(*bla*_NDM-1_) plasmids emerged more recently (earliest detections in 2015 and 2014, respectively), with varying frequencies each year (**Additional file 2: Fig. S4C, Fig. S4D**). These trends should be interpreted cautiously, however, as 25% of plasmids (n = 87) lacked collection date metadata and apparent decreases in plasmid counts in recent years likely reflects delays in sequence deposition. Nonetheless, these data show that the highly transmissible plasmids observed at our center are genetically conserved, despite being identified across diverse geographic regions.

## DISCUSSION

In this study, we characterized the diversity, prevalence, and genomic epidemiology of *bla*_NDM_-encoding plasmids within a single hospital system over a five-year period. We used both traditional genomic comparisons and epidemiologic methods to characterize plasmid transmission dynamics, and identified six clusters of genetically similar *bla*_NDM_-encoding plasmids which were repeatedly detected at our center. These plasmids differed in their genetic cargo, within-cluster similarity, detection frequency, and persistence over time. Plasmid clusters, and their nested bacterial host clusters, were found to have strong unit-level spatiotemporal associations. Additionally, we observed that highly abundant plasmids at our center were also prevalent globally, indicating both local and global dissemination of plasmids encoding carbapenem resistance.

The frequency of plasmid-encoded *bla*_NDM_ in our hospital system increased substantially beginning in April 2022. A report from the Centers for Disease Control and Prevention (CDC) indicated that the age-adjusted rate of *bla*_NDM_-encoding CRE increased by 461% across 29 states in the United States between 2019 and 2023 (35). Since expanded CRE surveillance began at our hospital after an outbreak investigation was initiated in April 2022, it is plausible that the increased prevalence of *bla*_NDM_-encoding CRE we observed may be at least partly attributable to increased detection of colonization following implementation of expanded perianal surveillance. However, the simultaneous national increase in *bla*_NDM_-encoding CRE during the same time period suggests that this pattern may also reflect a true rise in circulation of *bla*_NDM_-encoding plasmids.

Across a diverse set of *bla*_NDM_-encoding plasmid clusters, we observed a wide spectrum of similarity between plasmids, ranging from near-identical plasmid backbones (such as IncX3(*bla*_NDM-5_) and IncFIB(*bla*_NDM-1_)) to larger, more mosaic structures which retained similar resistance gene content alongside more variable accessory content (such as IncC and IncC/IncFIB(*bla*_NDM-1_), IncU(*bla*_NDM-1_), and IncHI2A(*bla*_NDM-1_)). For the highly conserved epidemic IncX3(*bla*_NDM-5_) plasmid, prior experimental studies have reported high plasmid stability, efficient conjugation, and minimal fitness cost in Enterobacterales—all features which could explain its success across a variety of host species (36). In contrast, broad-host-range IncC plasmids have been associated with large antibiotic resistance islands (including *bla*_NDM_-carriage) and frequent rearrangements (37), consistent with the more structurally variable IncC(*bla*_NDM-1_) plasmid architectures found in our dataset. Taken together, our results suggest that a small number of especially successful conjugative plasmid backbones were responsible for driving the dissemination of *bla*_NDM_ across species and patients at our center. Additionally, evolutionary processes like recombination and gene gain/loss are commonplace among resistance-encoding plasmids, complicating plasmid surveillance efforts in healthcare settings.

Our epidemiologic analysis showed that highly related *bla*_NDM_-encoding plasmid clusters, and nested host bacterial clusters, showed strong spatiotemporal associations with specific hospital units. These results suggest that shared unit-level exposures may be epidemiologically important for investigation of plasmid-associated outbreaks. Importantly, unit-level associations with plasmid clusters were found to be only partially explained by host isolate relatedness, supporting the hypothesis that both plasmid transmission and bacterial transmission contribute to multispecies *bla*_NDM_-encoding plasmid outbreaks, a dynamic which has been described previously (8,38). Unit-specific enrichment of plasmid clusters may be explained, in part, by environmental reservoirs present within hospital units (such as sink drains and plumbing biofilms), which have been shown to be colonized by multidrug-resistant Gram-negative bacteria harboring *bla*_NDM_-encoding plasmids over long periods (39). Thus, persistent unit-level signals may suggest unrecognized reservoirs which sustain ongoing exposures to plasmids, and could point to locations where environmental sampling and targeted intervention might be appropriate. Practically, these findings indicate that integration of WGS-based plasmid surveillance with epidemiologic data could assist with identification and prioritization of infection prevention measures such as staff education on *bla*_NDM_ carriage/transmission and enhanced environmental cleaning (8).

Through local and global comparisons, we identified the IncX3(*bla*_NDM-5_) and IncC(*bla*_NDM-1_) plasmid families as highly abundant and conserved, suggesting global dissemination. These plasmids encode type IV secretion systems permitting efficient conjugation across Enterobacterales (40–42). Further, studies investigating IncX3(*bla*_NDM-5_) properties have reported high plasmid retention, high conjugation frequencies, and low associated fitness cost (41,43). Such traits have likely contributed to the fact that this plasmid family is globally disseminated and has been identified from a wide variety of human, animal, and environmental sources (41,44–46). In contrast to the robust characterization of the IncX3(*bla*_NDM-5_) plasmid, there have been few prior reports of IncC(*bla*_NDM-1_)-associated outbreaks. A retrospective study described an IncC(*bla*_NDM-1_) outbreak in Rome, Italy where the plasmid additionally carried the 16S methyltransferase *rmt* gene conferring pan-aminoglycoside resistance, posing an increased public health threat (40). The clinically relevant resistances, diverse bacterial hosts, and global prevalence of this plasmid family suggest a need for further characterization and surveillance, as this plasmid appears poised for widespread dissemination.

This study had several limitations. First, WGS was limited to isolates identified through specific methods, including isolates either flagged by IP&C during suspected outbreaks, selected by EDS-HAT for surveillance, or submitted by the UPMC XDR Pathogens Lab. Second, we did not perform environmental sampling and could therefore not trace back a single source or reservoir for each of the plasmid clusters we detected. In addition, we were unable to account for healthcare encounters that occurred outside our hospital system, which limited our ability to fully assess spatiotemporal associations. Third, six patients lacked unit-level stay data, however missingness appeared to be random across the dataset. Fourth, unit-based active surveillance may have biased the results of our epidemiological analysis towards stronger unit-level associations; however, only one third of study isolates were collected through unit-based surveillance. Fifth, this study focused on shared spatial and temporal overlaps, and we recognize that these plasmids are likely propagated via a multitude of transmission routes, thus our findings are meant to be hypothesis generating. Further, we recognize that certain patient populations at risk for *bla*_NDM_ infection (*e.g.*, transplant patients) might be concentrated on certain high-risk units with increased CRE surveillance; thus, we recognize that some associations identified may simply reflect a concentration of at-risk patients in these units. Finally, our global plasmid analysis is likely biased towards countries with active CRE surveillance programs, such as China and North America. While metadata for publicly deposited sequences is often incomplete, we were still able to place our study plasmids within the global context and use the global dataset to identify avenues for future study.

## Conclusion

In conclusion, this study demonstrates the complex and dynamic nature of *bla*_NDM_-encoding plasmid dissemination within a tertiary-care hospital system over a multi-year period. We identified 11 distinct plasmid types, with transmission dominated by a small number of highly conserved plasmids, most notably IncX3(*bla*_NDM-5_) and IncC(*bla*_NDM-1_). These plasmids formed genetically related clusters that persisted over multiple years, encoded additional clinically relevant resistance genes, and demonstrated both plasmid-associated dissemination and bacterial expansion. Our results indicate that both plasmid clusters and their corresponding host bacterial clusters exhibit strong unit-level spatiotemporal associations within the hospital. Comparative genomic analysis with global plasmids showed several local plasmids belonged to widespread, genetically conserved plasmid backbones with distinct geographic and host-associated patterns. These findings emphasize the importance of continued plasmid-level surveillance to identify and intervene on transmission pathways with the ultimate goal of mitigating the spread of clinically relevant resistance genes in healthcare settings.

## DECLARATIONS

### Ethics approval and consent to participate

Ethics approval was obtained from the University of Pittsburgh Institutional Review Board (Protocol STUDY21040126).

### Consent for publication

Not applicable.

### Availability of data and materials

The datasets supporting the conclusions of this article are included within the article and its additional files. Sequencing data are publicly available through NCBI under BioProjects PRJNA981541 and PRJNA475751. All associated accession numbers are listed in the supplementary information files.

### Competing interests

LHH serves on the scientific advisory board of Next Gen Diagnostics. AJS is a consultant for Next Gen Diagnostics. RKS serves as a consultant to bioMérieux and Informuta. All other authors report no conflicts of interest.

## Funding

This work is funded in part by the National Institute of Allergy and Infectious Diseases, National Institutes of Health (NIH) grants R01AI127472 (Enhanced Detection System for Healthcare-Associated Transmission of Infection) and R21AI178369 (Tracking plasmid spread and transmission in the hospital: A novel tool for infection prevention and control). The NIH played no role in data collection, analysis, or interpretation; study design; writing of the manuscript; or decision to submit for publication.

## Authors’ contributions

NJR, EMM, and DVT conceived and designed the study. NJR and EMM curated the dataset and conducted the investigation, including formal epidemiologic and bioinformatic analyses. NJR and EMM also performed data validation, created the visualizations, and wrote the original draft and subsequent revisions. SB contributed to formal analysis and visualizations for the global plasmid similarity analysis and manuscript revision. MPG contributed to data curation, including uploading WGS data to public repositories, and manuscript revision. KS was responsible for the collection and curation of epidemiologic data. KDW, LLP, and AJS contributed to acquisition of funding, laboratory resources for sequencing, and manuscript revision. RKS contributed collection and characterization of CRE isolates used in the study, and manuscript revision. GMS contributed to supervision, funding acquisition, and manuscript revision, as well as providing IP&C expertise. DVT and LHH jointly provided overall leadership and supervision; contributed to conceptualization, resources, funding acquisition, and manuscript writing and revision. DVT additionally supported project administration. All authors read and approved the final manuscript.

## Acknowledgements.

Special thanks to Yohei Doi, MD, PhD, for providing select samples included in the study, Jane Marsh, PhD, for her role in developing the EDS-HAT infrastructure, and Vaughn Cooper, PhD, for whole genome sequencing assistance.

## Supplementary Information

**Additional file 1: Table S1-S3.** Table S1: Sample metadata and plasmid-encode genetic features. Table S2: Unit-level odds ratios for plasmid and bacterial host clusters based on qualifying shared-exposure stay pairs. Table S3: Global plasmid metadata and presence of antimicrobial resistance genes.

**Additional file 2: Figures S1-S4.** Figure S1: Histograms of pairwise host isolate SNPs and plasmid containment / DCJ-Indel distances. Figure S2: Qualifying shared-exposure stay pairs. Figure S3: Alignment of IncX3 (*bla*_NDM-5_) plasmids. Figure S4. Temporal distribution of global and local *bla*_NDM_-positive plasmids by plasmid family and bacterial species.

**Additional file 3: Extended Data.** Raw data for epidemiological analyses (‘All Stay Pairs’, ‘Stay Pairs Collapsed’) and genomic analyses (‘Containment and DCJ’, ‘Isolate SKA SNPs’, ‘IncC IncFIB Addtl Plasmids’).

---

**Figure S1:**
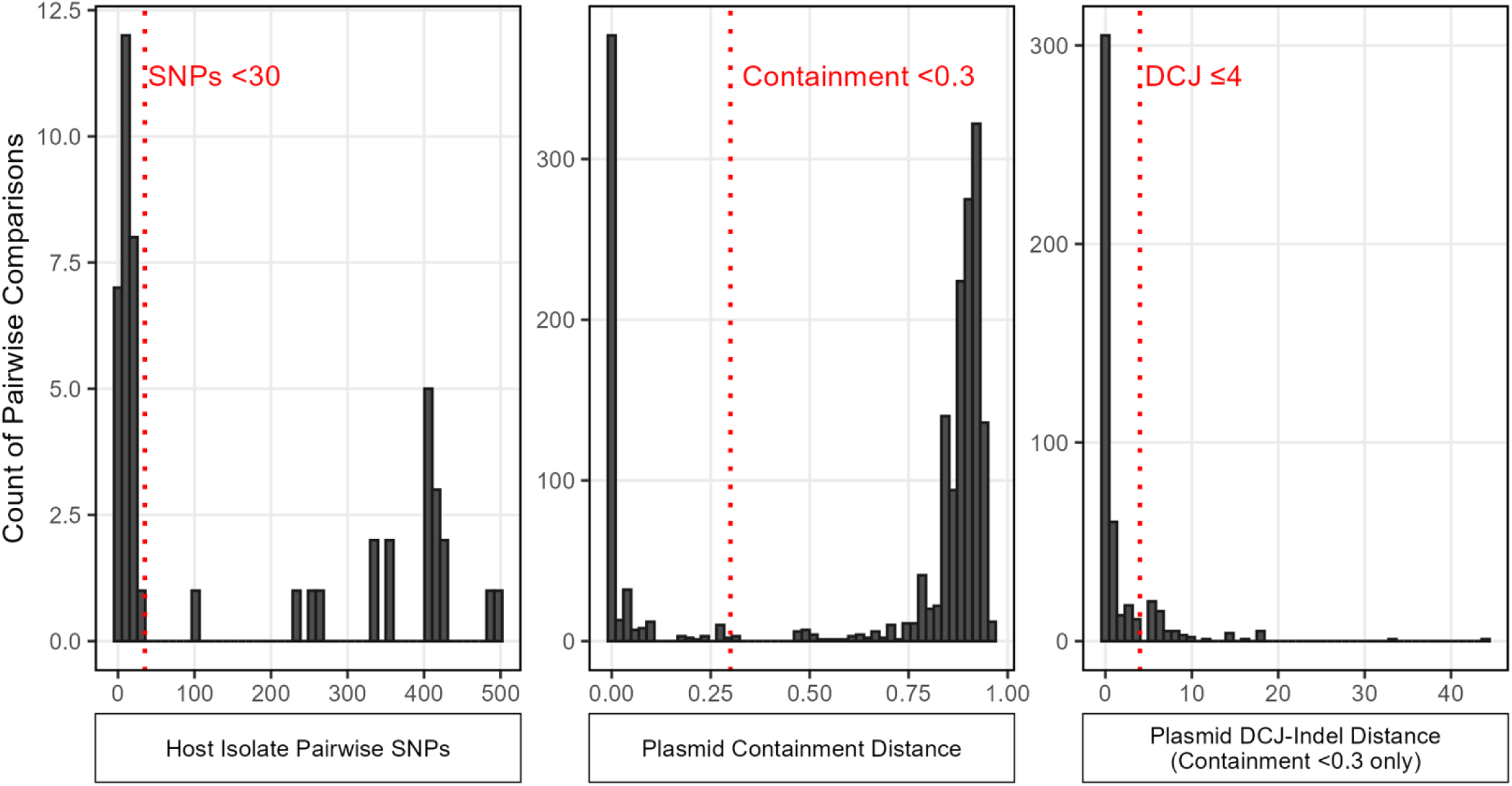
Histograms of pairwise host isolate SNPs and plasmid containment / DCJ-Indel distances. The distribution of pairwise SNPs (from SKA, truncated to show comparisons ≤500 SNPs) calculated between study isolates of the same species and multi-locus sequence type are shown on the leftmost panel; the threshold for host relatedness (≤30 SNPs) is shown as a dotted red line. Pairwise containment and DCJ-Indel distances calculated between study plasmids are shown in the center and right panels, respectively; thresholds for containment (<0.3) and DCJ-Indel (≤4) distances are shown with dotted red lines. DCJ-Indel distances were calculated only between plasmids with containment distances <0.3.

**Figure S2:**
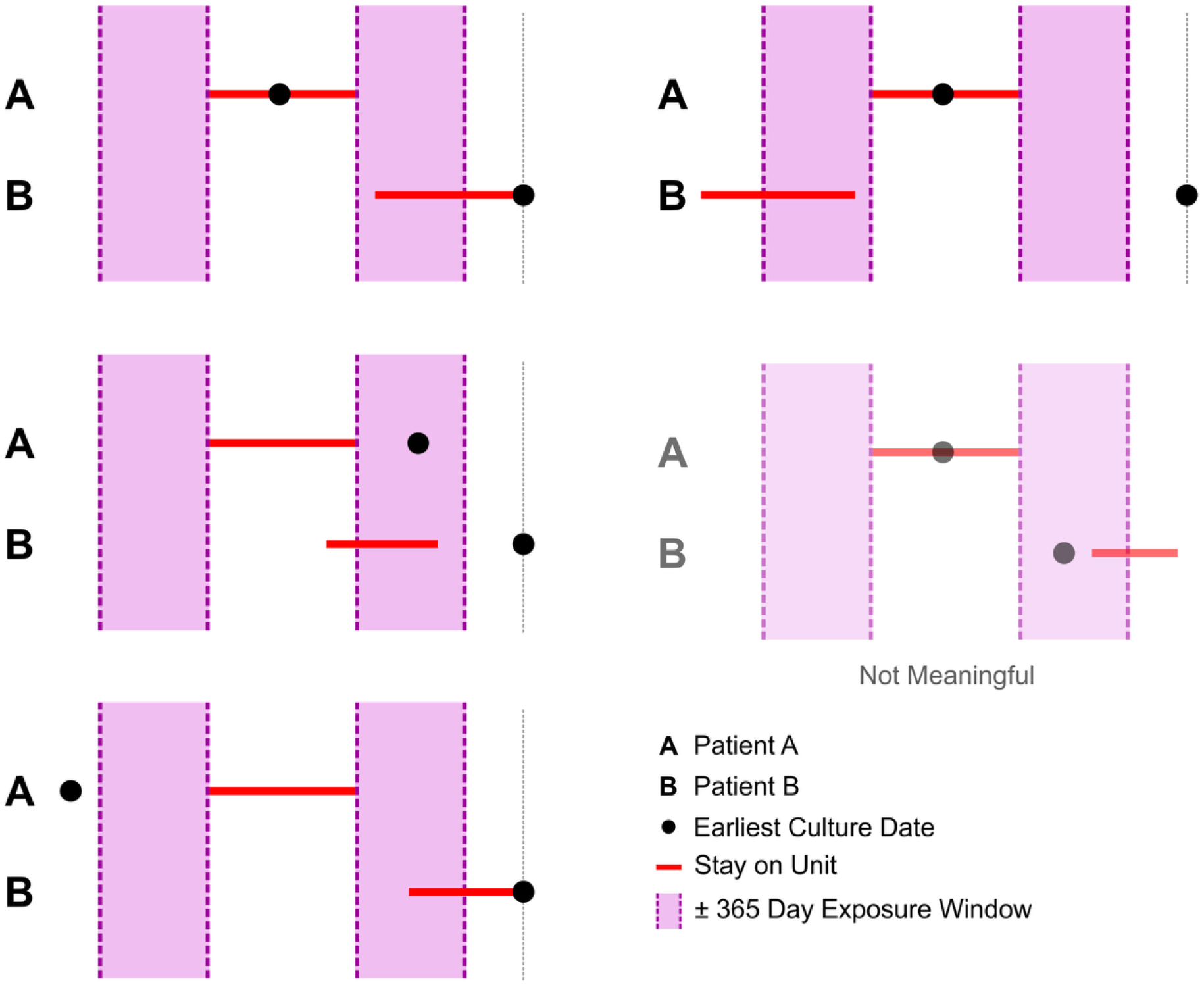
Qualifying shared-exposure stay pairs. Qualifying shared-exposure stay pairs between example Patients A and B are shown. For each pair, the patient with the earliest positive culture date (denoted by black circles) was labeled Patient A. A ±365-day exposure window (shaded pink) was extended from the admission and discharge dates for Patient ’s stay (red lines indicate unit stays). Pairs were considered a qualifying shared-exposure pair when Patient ’s stay intersected the exposure window and ended on or before Patient ’s earliest positive culture date. Bottom right panel illustrates an example of a non-qualifying stay pair.

**Figure S3:**
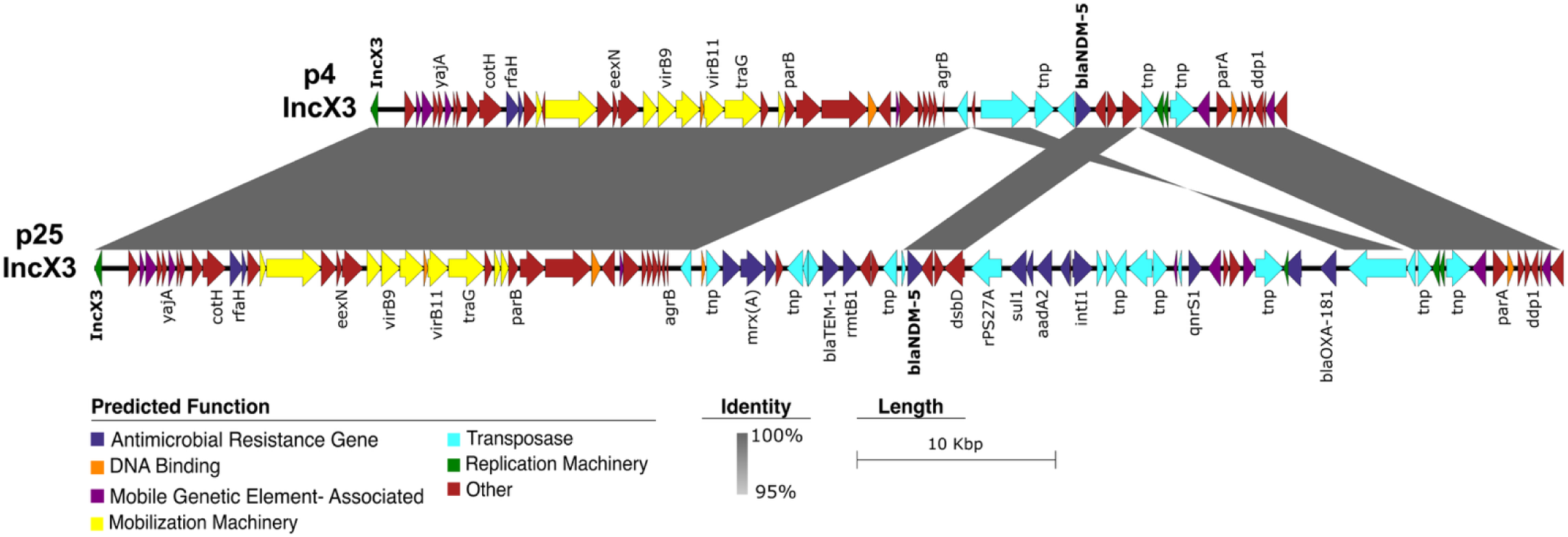
Alignment of IncX3(*bla*_NDM-5_) plasmids. Nucleotide sequence alignment between IncX3(*bla*_NDM-5_) plasmids p4 (46 kb) and p25 (73 kb) were visualized using EasyFig v2.2.2. Genes were annotated using Bakta v1.9.2 and were grouped into predicted functional categories as indicated. Nucleotide identity, calculated from BLASTn comparisons using a 1,000-bp window size, is shown from 95% (light grey) to 100% (dark grey).

**Figure S4.**
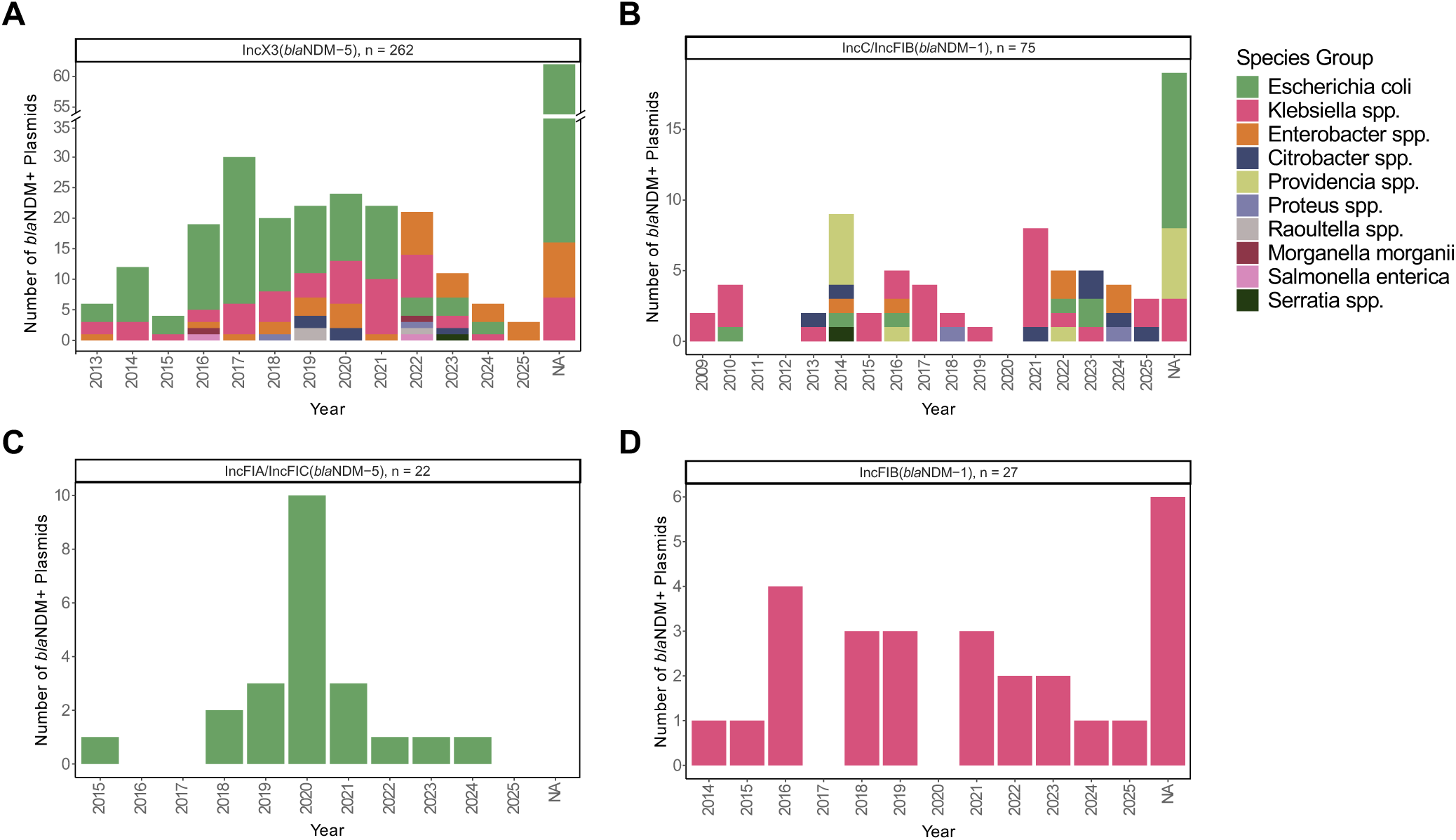
Temporal distribution of global and local *bla*_NDM_-encoding plasmids by plasmid family and bacterial species. Stacked bar plots showing the yearly distribution of *bla*_NDM_-positive plasmids, stratified by plasmid family and bacterial species. Panels correspond to **(A)** IncX3(*bla*_NDM-5_), **(B)** IncC/IncFIB(*bla*_NDM-1_), **(C)** IncFIA/FIC(*bla*_NDM-5_), and **(D)** IncFIB(*bla*_NDM-1_) plasmid families. The x-axis represents year of isolation, and the y-axis indicates the number of plasmids identified each year. Bars are colored by species group, as indicated in the legend.

